# Effect of Heparin and Tocilizumab in Patients with Severe COVID-19: The HEPMAB Randomized Clinical Trial

**DOI:** 10.1101/2023.12.22.23300466

**Authors:** Lucas Trindade Cantú Ribeiro, Lucas Trindade Cantú Ribeiro, Giovanni Landoni, Vinícius Caldeira Quintão, Marcus Vinicius Guimaraes de Lacerda, Stephanie Itala Rizk, Isabela Bispo Santos da Silva Costa, Fernanda Thereza de Almeida Andrade, Edielle de Sant’Anna Melo, Nestor Cordeiro dos Santos Neto, Thalita Barbosa de González, Laisse Barreto Ferreira Reis, Valmir de Freitas Costa, Theuran Inahja Vicente Machado, Sabrina Corrêa da Costa Ribeiro, Alexandra Patricia Zilli Vieira, RN Natassja Huemer, Fabricio Silva, Clarice Lee Park, Júlia Tizue Fukushima, Milton Henrique Guimarães Junior, Marcos de Abreu Lima Cota, Lucas Tokio Kawahara, Cecília Chie Sakaguchi Barros, Alicia Muller, Leticia Nakada, Bruno Fioravanti de Paiva Pinto, Fabrício Sanchez Bergamin, RN Eduesley Santana Santos, Fernando Augusto Marinho dos Santos Figueira, Leonardo Jorge Cordeiro de Paula, Roberto Kalil Filho, Ludhmila Abrahão Hajjar

## Abstract

**Background:** Clinical presentation of severe Coronavirus disease 2019 (COVID-19) is associated to an intense inflammatory response and thrombogenesis. The benefits of the association of interleukin-6 receptor blockade (tocilizumab) and therapeutic-dose anticoagulation remains unclear. We aimed to assess whether heparin and tocilizumab could effectively reduce inflammation and thrombogenesis in severe COVID-19 patients.

**Methods:** This is an open-label, multicenter, randomized, clinical trial, involving patients with severe COVID-19 infection. Eligible patients were randomly assigned in a 1:1:1:1 ratio to receive either therapeutic or prophylactic anticoagulation with heparin, with or without an intravenous single dose of tocilizumab. The participants in the study were assigned to one of the four distinct arms: 1) therapeutic anticoagulation; 2) prophylactic anticoagulation; 3) therapeutic anticoagulation plus a single intravenous dose of tocilizumab; and 4) prophylactic anticoagulation plus a single intravenous dose of tocilizumab. The primary outcome was clinical improvement at day 30, defined as a composite of hospital discharge and/or a reduction of at least 2 points of the modified ordinal scale of 7 points recommended by the World Health Organization.

**Results:** We enrolled 308 patients. Patients randomized to receive therapeutic anticoagulation more frequently had clinical improvement at day 30 when compared to the prophylactic anticoagulation patients [64/75 (85%) versus 51/80 (64%), odds ratio, 3.31; 95% confidence interval, 1.51; 7.26 P=0.003]. Major bleeding was more frequent in the therapeutic anticoagulation group (6.7%) and in the therapeutic anticoagulation plus tocilizumab group (5.0%), compared to the prophylactic anticoagulation group (P=0.02). All-cause mortality at day 30 was significantly lower in therapeutic anticoagulation group (9.3%), when compared to prophylactic anticoagulation group (28.7%), therapeutic anticoagulation plus tocilizumab group (21.5%) and prophylactic anticoagulation plus tocilizumab group (25.7%), P=0.02.

**Conclusions:** In this randomized clinical trial involving severe COVID-19 patients, therapeutic anticoagulation resulted in clinical improvement at 30 days. Even if therapeutic anticoagulation increased bleeding, it was associated with a reduced overall mortality. Tocilizumab did not provide additional benefits to heparin in COVID-19 patients.

**Trial registration:** Clinicaltrials.gov NCT04600141. Registered October 22, 2020. https://www.clinicaltrials.gov/study/NCT04600141?term=NCT04600141&rank=1

## Background

Coronavirus disease 2019 (COVID-2019) pandemic has resulted in over 704,000 fatalities and more than 37 million confirmed cases in Brazil.[1] This number reflect the high burden of disease in a country with 214 million habitants with regional disparities within the health system, lack of a high-quality care for critically ill patients, and scarcity of intensive care beds.[2]

COVID-19 is characterized by an initial phase of viral replication, followed by a second phase, resulting in different phenotypes of disease, accordingly to the patient’s immune system response.[3] The severe form is characterized by an intense inflammatory response, driven by T-lymphocytes, cytokine release, and marked by endothelial activation and thrombogenesis.[4] Antiviral, anticoagulants, and immunomodulatory medications have been the mainstay of treatment in critical patients, with demonstrated benefits from remdesivir and steroids.[5-8]

Administering anticoagulation is crucial for managing severe cases of COVID-19, but the optimal dosage, type of anticoagulant, and duration of treatment remain a topic of debate.[9-14] Recent published data have shown that in severe disease, therapeutic doses of heparin reduce complications, while in critically ill patients, there is no evidence for benefit.^15,16^

Tocilizumab is an interleukin 6 (IL-6) receptor antagonist drug, primarily used to treat autoimmune diseases, such as rheumatoid arthritis, systemic juvenile idiopathic arthritis, and cytokine release syndrome after CAR-T-Cells therapy.[15] Retrospective data, published at the onset of the pandemics, suggested that tocilizumab reduces death and rates of mechanical ventilation in SARS-CoV-2 patients needing high oxygen support.[16] Published randomized clinical trials have yielded inconsistent findings regarding the effectiveness of tocilizumab in treating COVID-19 patients.[17-24] The data from the Recovery trial[25], an important publication that has led to changes in treatment guidelines for COVID-19[26], showed a marginal benefit of tocilizumab in lowering mortality rates, and the reduction in the risk of death was not statistically significant for all patient subgroups. Then, we hypothesized that the early association of anticoagulation with immunomodulation therapies could improve outcomes and survival rates in these patients.

Therefore, this study aimed to assess whether heparin and tocilizumab could effectively reduce inflammation and thrombogenesis in severe COVID-19 patients.

## Methods

### Trial Design

A randomized, controlled, open-label, multicenter, parallel, pragmatic trial was conducted in eight high complexity COVID-19 referral hospitals in Brazil. The protocol was registered in Clinicaltrials.gov (NCT04600141). The trial was designed by the executive committee (see the Supplementary Appendix), and protocol and informed consent were approved by the ethics committees of Hospital das Clínicas, Faculdade de Medicina, Universidade de São Paulo, the coordinator center, and by each participating sites (Table e1, Supplementary Appendix).

The trial was overseen by a safety monitoring committee. The initial version of the manuscript was drafted by the first and last authors, developed by the writing committee, and approved by all members of the trial steering committee. The executive committee vouches for the completeness and accuracy of the data and for the fidelity of the trial to the Consolidated Standards of Reporting Trials.

### Participants

We included individuals aged ≥ 18 who had been hospitalized with a confirmed diagnosis of COVID-19 (by real-time polymerase chain reaction – RT-PCR) and were actively screened by the trial team, within 10 days from symptoms onset, and/or radiologic evidence of disease confirmed by chest radiography or tomography and needing at least 4 L/min of oxygen to keep oxygen saturation ≥ 93%. Exclusion criteria were patients with a high risk of bleeding, known or suspected adverse reaction to unfractionated heparin (UFH) or tocilizumab, active thrombosis requiring anticoagulants, thrombolytic therapy in the last 3 days, use of glycoprotein IIb/IIIa inhibitors within the previous 7 days and septic shock. Complete information on inclusion and exclusion criteria is provided in the Supplementary Appendix.

### Randomization and masking

Eligible patients were randomly assigned in a 1:1:1:1 ratio. Randomization was stratified according to the trial site at enrollment using a Web-based system.

Patients and their health care providers were aware of treatment administered, while the statisticians analyzing the data were kept blinded to the treatment status.

### Procedures

After written informed consent, data was continuously entered into the web-based database. Patients received standard supportive care at the trial site hospital. If a hospital had a written policy for COVID-19 treatment, patients could receive corticosteroids, remdesivir, antimicrobial therapy for bacterial or fungal infections, and prophylaxis for *Strongyloides*. Concomitant experimental drugs were not allowed. The participants in the study were assigned to one of the four distinct arms: 1) therapeutic anticoagulation (intravenous UFH to keep aPTT at 50-70s or subcutaneous enoxaparin 1 mg/Kg twice a day till hospital discharge); 2) prophylactic anticoagulation (subcutaneous UFH 5,000 IU three times a day or subcutaneous enoxaparin 40 mg once a day till hospital discharge); 3) therapeutic anticoagulation plus a single intravenous dose of tocilizumab 8 mg/Kg; and 4) prophylactic anticoagulation plus a single intravenous dose of tocilizumab 8 mg/Kg.

Patients were evaluated daily until death, discharge, or day 30 after randomization. Discharged patients were followed by a telephone call on day 30.

Dosage of IL-6, D-dimer, troponin, C-reactive protein (CRP), N-terminal pro B-type natriuretic peptide (NT-proBNP, and ferritin were performed immediately after randomization, in 72 hours (D3) and 7 days after the start of treatment (D7) (see Supplementary Appendix).

### Outcomes

Primary outcome was the proportion of patients with clinical improvement at day 30, defined by hospital discharge or a reduction of at least 2 points compared to baseline on the modified ordinal scale of 7 points recommended by the WHO. (Table e2, Supplementary Appendix).

Secondary outcomes included the proportion of participants who needed invasive mechanical ventilation, duration of invasive mechanical ventilation, proportion of participants who needed vasopressor, as well as the duration of their vasopressor use, renal failure by the Acute Kidney Injury Network (AKIN) criteria, cardiovascular complications, venous thromboembolism, 30-day mortality, intensive care unit (ICU) length of stay, and duration of hospitalization.

Safety outcomes assessed comprised of serious adverse events, defined as major bleeding, heparin-induced thrombocytopenia, and septic shock. Biomarkers levels were also compared among groups.

### Sample size and statistical analysis

The study was designed to achieve an 80% power for detecting an increase in the proportion of patients experiencing clinical improvement from 55% in the comparator groups to 80% in the therapeutic anticoagulation plus tocilizumab group, considering a 2-sided type I error of 5%. Based on these assumptions, and considering 10% loss of follow-up, the estimated sample size was 308 patients divided in 4 arms. The hypothesis of a 55% occurrence of the primary end point in the three comparator arms was derived from preliminary data of a Hospital das Clínicas, Faculdade de Medicina, Universidade de São Paulo COVID-19 patient cohort. The sample size was adjusted based on Bonferroni multiple comparisons.

Primary efficacy was conducted as intention-to-treat (ITT) analysis. During the four weeks after randomization, the Chi-square test in an asymptotic form and the odds ratio with its two-tailed 95% confidence interval (CI) were used to compare the percentage of patients with deterioration in their medical state among the four arms.

Kaplan-Meier curves and Cox proportional hazard model were used for unadjusted analysis to 30-day mortality. For the secondary outcome of 30-day mortality, the log-rank observed minus expected statistic and its variance were used to test the null hypothesis of equal survival curves and to calculate the one-step estimate of the average mortality rate ratio.

Quantitative variables were expressed as mean and standard deviation and compared between groups with one-way ANOVA and Bonferroni correction was used to account for multiple comparisons. For non-normally distributed variables, the Kruskal-Wallis test followed by Dunn’s post hoc test was used to compare the groups. The categorical data were assessed using Chi-square or the likelihood ratio test.

We reported the P value efficacy analysis (two-tailed; significance defined as P < .05). Statistical analyses were performed with SPSS, version 25 (IBM Corp. Released 2017, Armonk, NY).

## Results

### Characteristics of patients

From November 16, 2020, through August 02, 2021, a total of 702 patients were assessed for eligibility. Of these patients, 310 underwent randomization, 2 withdrew consent and 308 were included in the intention-to-treat analysis (Figure 1).

**Figure 1.**
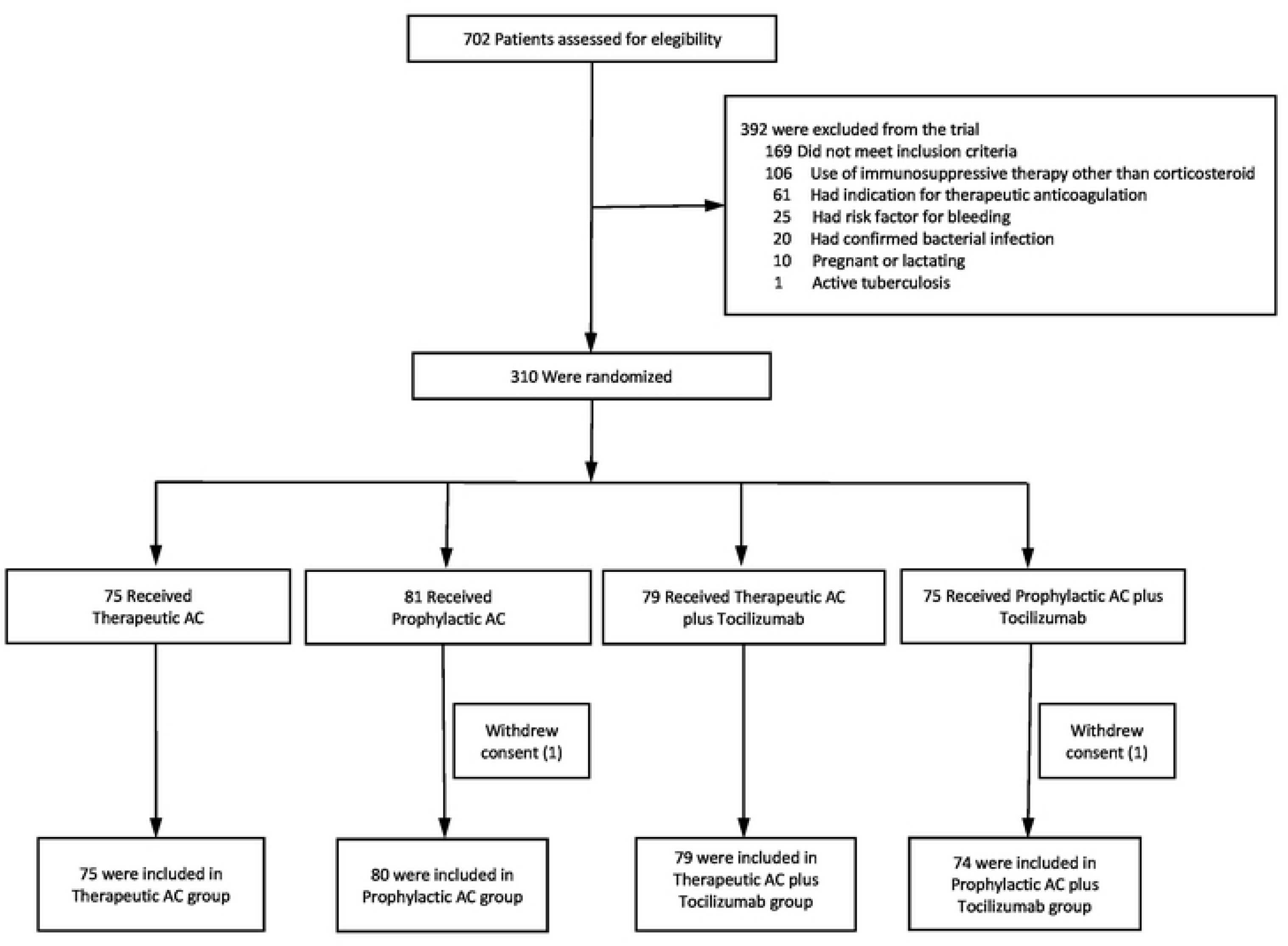
Study flow diagram. Screening, enrollment, randomization, and inclusion in analysis. AC: anticoagulation.

The baseline characteristics and comorbidities of the patients (Table 1) were well balanced among groups. Mean (±SD) age was 53 ± 13 years, 183 patients (59%) were male, and 63% of patients were white. Most patients required high-flow oxygen therapy (191, 62%), 49 (16%) needed non-invasive mechanical ventilation, and 68 (22%) required invasive mechanic ventilation. The median concentration of CRP was 110 mg/L (IQR, 64.9 to 175.3); ferritin was 708 ng/mL (IQR, 411 to 1225); D-dimer was 884 ng/mL(IQR, 527 to 1730); lactate dehydrogenase (LDH) was 340 U/L(IQR, 289 to 413) and IL-6 was 18.4 ng/mL (IQR, 7 to 52) (Table e3, Supplementary Appendix).

**Table 1.**
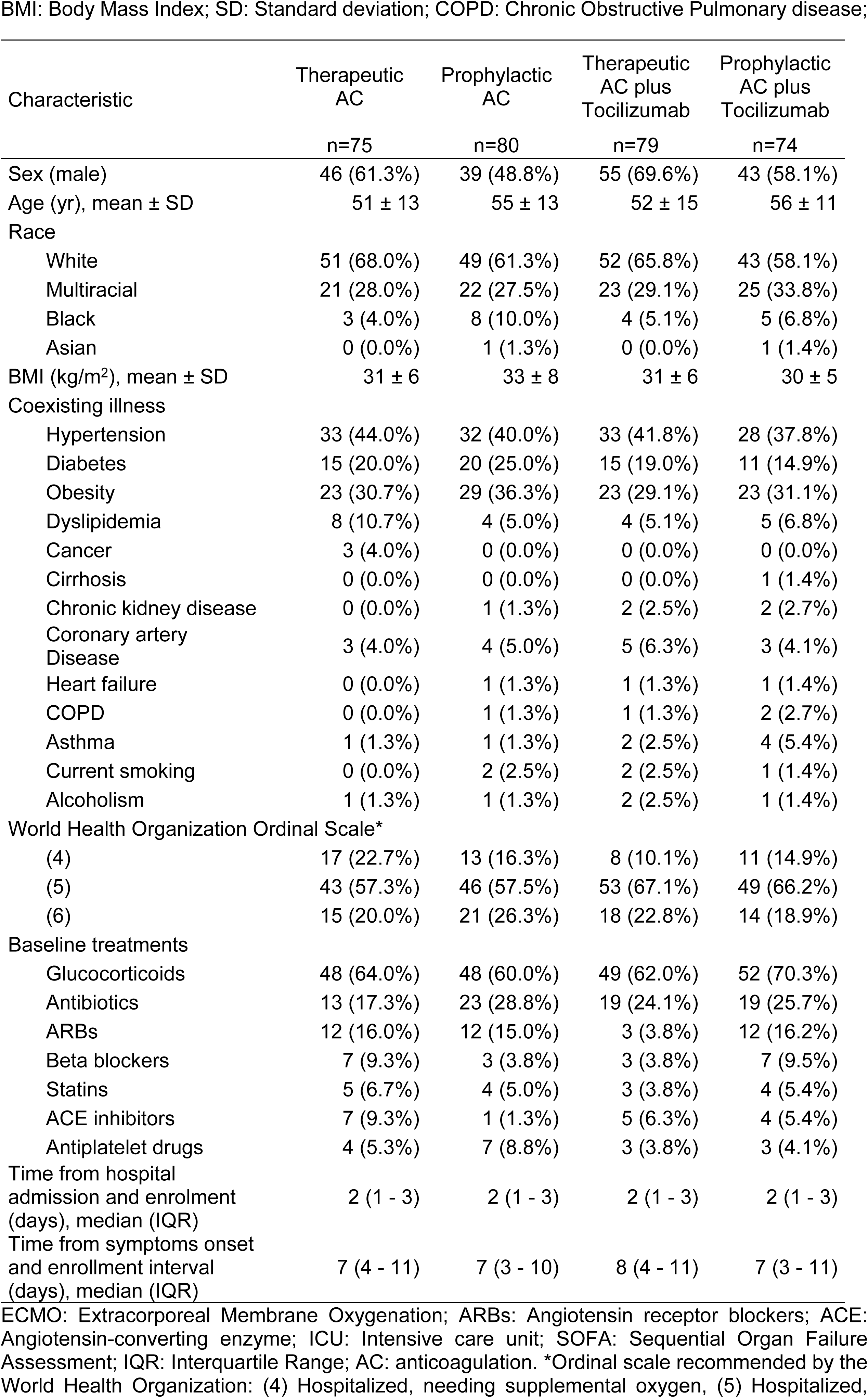
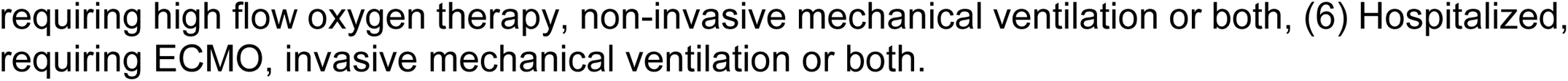
Baseline characteristics of patients.

BMI: Body Mass Index; SD: Standard deviation; COPD: Chronic Obstructive Pulmonary disease;
| Characteristic | Therapeutic<br>AC<br>n=75 | Prophylactic<br>AC<br>n=80 | Therapeutic<br>AC plus<br>Tocilizumab<br>n=79 | Prophylactic<br>AC plus<br>Tocilizumab<br>n=74 |
| --- | --- | --- | --- | --- |
| Sex (male) | 46 (61.3%) | 39 (48.8%) | 55 (69.6%) | 43 (58.1%) |
| Age (yr), mean ± SD | 51 ± 13 | 55 ± 13 | 52 ± 15 | 56 ± 11 |
| Race |  |  |  |  |
| White | 51 (68.0%) | 49 (61.3%) | 52 (65.8%) | 43 (58.1%) |
| Multiracial | 21 (28.0%) | 22 (27.5%) | 23 (29.1%) | 25 (33.8%) |
| Black | 3 (4.0%) | 8 (10.0%) | 4 (5.1%) | 5 (6.8%) |
| Asian | 0 (0.0%) | 1 (1.3%) | 0 (0.0%) | 1 (1.4%) |
| BMI (kg/m<sup>2</sup>), mean ± SD | 31 ± 6 | 33 ± 8 | 31 ± 6 | 30 ± 5 |
| Coexisting illness |  |  |  |  |
| Hypertension | 33 (44.0%) | 32 (40.0%) | 33 (41.8%) | 28 (37.8%) |
| Diabetes | 15 (20.0%) | 20 (25.0%) | 15 (19.0%) | 11 (14.9%) |
| Obesity | 23 (30.7%) | 29 (36.3%) | 23 (29.1%) | 23 (31.1%) |
| Dyslipidemia | 8 (10.7%) | 4 (5.0%) | 4 (5.1%) | 5 (6.8%) |
| Cancer | 3 (4.0%) | 0 (0.0%) | 0 (0.0%) | 0 (0.0%) |
| Cirrhosis | 0 (0.0%) | 0 (0.0%) | 0 (0.0%) | 1 (1.4%) |
| Chronic kidney disease | 0 (0.0%) | 1 (1.3%) | 2 (2.5%) | 2 (2.7%) |
| Coronary artery<br>Disease | 3 (4.0%) | 4 (5.0%) | 5 (6.3%) | 3 (4.1%) |
| Heart failure | 0 (0.0%) | 1 (1.3%) | 1 (1.3%) | 1 (1.4%) |
| COPD | 0 (0.0%) | 1 (1.3%) | 1 (1.3%) | 2 (2.7%) |
| Asthma | 1 (1.3%) | 1 (1.3%) | 2 (2.5%) | 4 (5.4%) |
| Current smoking | 0 (0.0%) | 2 (2.5%) | 2 (2.5%) | 1 (1.4%) |
| Alcoholism | 1 (1.3%) | 1 (1.3%) | 2 (2.5%) | 1 (1.4%) |
| World Health Organization Ordinal Scale* |  |  |  |  |
| (4) | 17 (22.7%) | 13 (16.3%) | 8 (10.1%) | 11 (14.9%) |
| (5) | 43 (57.3%) | 46 (57.5%) | 53 (67.1%) | 49 (66.2%) |
| (6) | 15 (20.0%) | 21 (26.3%) | 18 (22.8%) | 14 (18.9%) |
| Baseline treatments |  |  |  |  |
| Glucocorticoids | 48 (64.0%) | 48 (60.0%) | 49 (62.0%) | 52 (70.3%) |
| Antibiotics | 13 (17.3%) | 23 (28.8%) | 19 (24.1%) | 19 (25.7%) |
| ARBs | 12 (16.0%) | 12 (15.0%) | 3 (3.8%) | 12 (16.2%) |
| Beta blockers | 7 (9.3%) | 3 (3.8%) | 3 (3.8%) | 7 (9.5%) |
| Statins | 5 (6.7%) | 4 (5.0%) | 3 (3.8%) | 4 (5.4%) |
| ACE inhibitors | 7 (9.3%) | 1 (1.3%) | 5 (6.3%) | 4 (5.4%) |
| Antiplatelet drugs | 4 (5.3%) | 7 (8.8%) | 3 (3.8%) | 3 (4.1%) |
| Time from hospital<br>admission and enrolment<br>(days), median (IQR) | 2 (1 - 3) | 2 (1 - 3) | 2 (1 - 3) | 2 (1 - 3) |
| Time from symptoms onset<br>and enrollment interval<br>(days), median (IQR) | 7 (4 - 11) | 7 (3 - 10) | 8 (4 - 11) | 7 (3 - 11) |

ECMO: Extracorporeal Membrane Oxygenation; ARBs: Angiotensin receptor blockers; ACE: Angiotensin-converting enzyme; ICU: Intensive care unit; SOFA: Sequential Organ Failure Assessment; IQR: Interquartile Range; AC: anticoagulation. \*Ordinal scale recommended by the World Health Organization: (4) Hospitalized, needing supplemental oxygen, (5) Hospitalized,
requiring high flow oxygen therapy, non-invasive mechanical ventilation or both, (6) Hospitalized, requiring ECMO, invasive mechanical ventilation or both.

Patients were randomized after 7 (IQR, 4 – 10) days of symptoms onset and within 2 (IQR, 1 – 3) days from hospital admission. The majority of patients were given glucocorticoids (197, 64%). Signs and symptoms, vital signs and concomitant treatments at baseline are provided in the Table e4, Table e5 and Table e6 (Supplementary Appendix).

All 153 patients randomized to groups 3 and 4 received an intravenous dose of tocilizumab. Heparin protocol deviations are presented in Table e7 (Supplementary Appendix).

### Primary outcome

Patients in the therapeutic anticoagulation group more frequently reached the clinical improvement endpoint when compared to the reference group (prophylactic anticoagulation): 64/75 (85%) versus 51/80 (64%), odds ratio, 3.31; 95% confidence interval [CI], 1.51; 7.26 P=0.003. (Table 2)

**Table 2.** Outcomes.

| Outcomes | Therapeutic AC<br>n=75 | Prophylactic AC<br>n=80 | Therapeutic AC +<br>Tocilizumab<br>n=79 | Prophylactic AC +<br>Tocilizumab<br>n=74 | P |
| --- | --- | --- | --- | --- | --- |
| **Primary outcome** |  |  |  |  |  |
| Hospital discharge or reduction of at least 2 points on the ordinal scale at Day 30, No. (%) | 64 (85) | 51 (64) | 53 (67) | 49 (66) | .013<sup>a</sup> |
| *Odds ratio (95%CI)* | *3.31 (1.51-7.26)* | *Reference* | *1.16 (0.60-2.23)* | *1.12 (0.57-2.16)* |  |
| *P* | *.003* |  | *.66* | *.57* |  |
| **Secondary Outcomes** |  |  |  |  |  |
| 30-day mortality, No. (%) | 7 (9) | 23 (29) | 17 (22) | 19 (26) | .020<sup>a</sup> |
| *Hazard ratio (95% CI)* | *0.30 (0.13-0.69)* | *Reference* | *0.75 (0.40-1.40)* | *0.89 (0.49-1.64)* |  |
| *P* | *.005* |  | *.36* | *.71* |  |
| ICU length of stay, median (IQR), days | 11 (5-27) | 14 (6-25) | 17 (7-24) | 16 (9-26) | .87<sup>c</sup> |
| Patients requiring invasive mechanical ventilation | 12 (16) | 13 (16) | 14 (18) | 16 (22) | .79<sup>a</sup> |
| Days of invasive mechanical ventilation, median (IQR) | 7 (2-16) | 10 (7-14) | 4 (2-11) | 4 (2-8) | .11<sup>c</sup> |
| Patients requiring vasopressors, No. (%) | 12 (16) | 12 (15) | 12 (15) | 14 (19) | .91<sup>a</sup> |
| Duration of vasopressors, median (IQR), days | 7 (2-20) | 10 (8-14) | 3 (1-16) | 4 (2-8) | .10<sup>c</sup> |
| Acute Kidney Injury (AKIN≥1) , No. (%) | 46 (61) | 47 (59) | 45 (57) | 37 (50) | .54<sup>a</sup> |
| End-stage renal disease, No. (%) | 4 (5) | 5 (6) | 5 (6) | 1 (1) | .33<sup>b</sup> |
| Cardiovascular outcomes, No. (%): |  |  |  |  |  |
| - Myocardial injury | 10 (13) | 8 (10) | 7 (9) | 9 (12) | .81<sup>a</sup> |
| - Arrhythmias | 11 (15) | 8 (10) | 7 (9) | 12 (16) | .44<sup>a</sup> |
| - Acute myocardial infarction | 0 (0) | 0 (0) | 0 (0) | 1 (1) | .41<sup>b</sup> |
| - Cardiogenic shock | 0 (0) | 1 (1) | 0 (0) | 1 (1) | .43<sup>b</sup> |
| - Myocarditis/pericarditis | 0 (0) | 2 (3) | 1 (1) | 0 (0) | .23<sup>b</sup> |
| - Ventricular dysfunction | 1 (1) | 2 (3) | 1 (1) | 1 (1) | .92<sup>b</sup> |
| Venous thromboembolism, No. (%) | 0 (0) | 3 (4) | 1 (1) | 4 (3) | .08<sup>b</sup> |

a: Pearson's chi-squared test; b: likelihood-ratio test; c: Kruskal-Wallis test. AC: anticoagulation; IQR: Interquartile Range; CI: Confidence Interval.

### Secondary outcomes

Mortality at day 30 was significantly lower in therapeutic anticoagulation group (9.3%), when compared to prophylactic anticoagulation (28.7%), therapeutic anticoagulation plus tocilizumab (21.5%) and prophylactic anticoagulation plus tocilizumab groups (25.7%), P=0.02 (Figure 2). Causes of death are provided in Table e8 (Supplementary Appendix). No difference among groups was found in other secondary outcomes. (Table 2)

**Figure 2.**
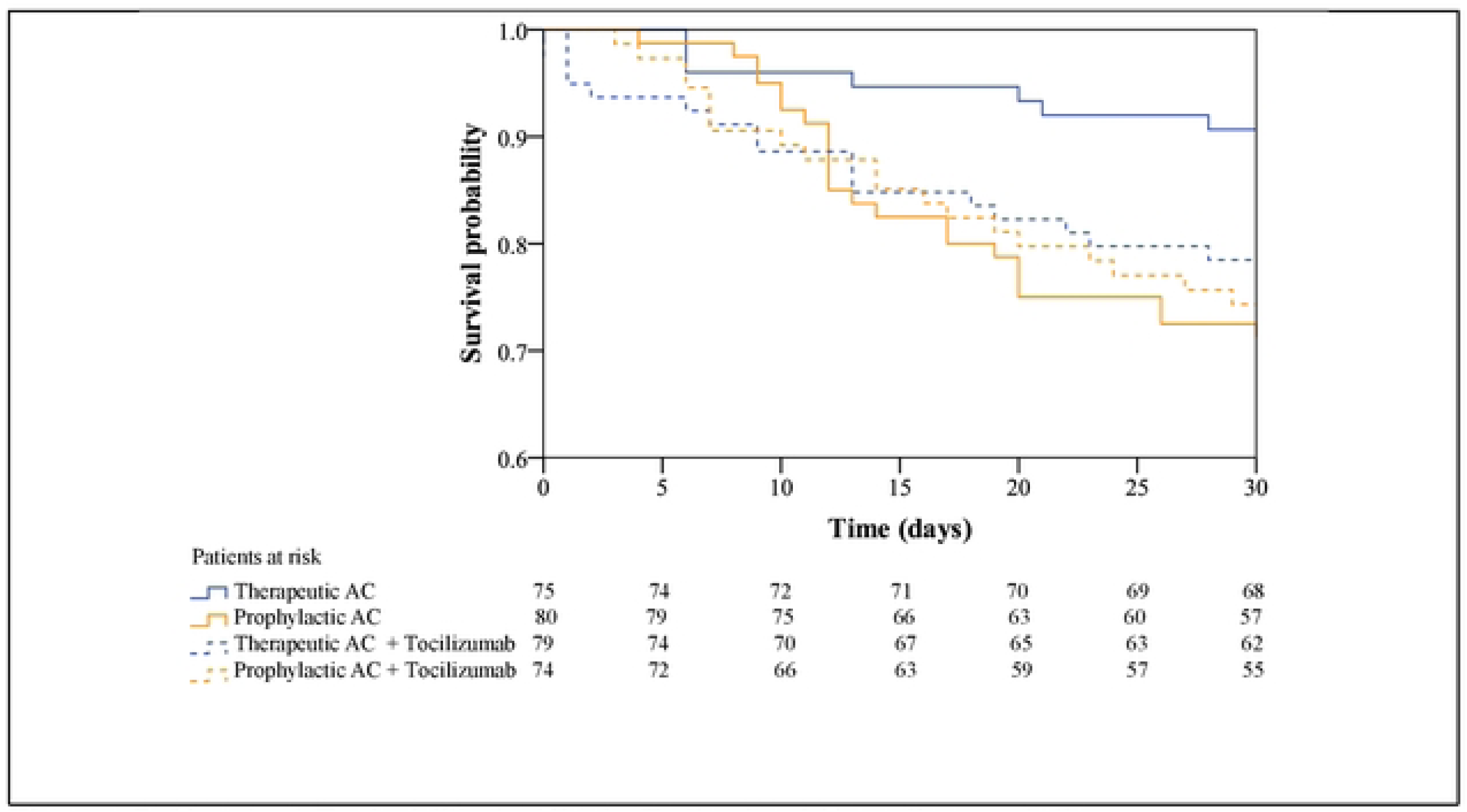
Survival Probability in Study Patients. AC: anticoagulation.

Therapeutic heparin administration significantly reduced D-dimer levels while tocilizumab significantly reduced CRP during the first 7 days of follow-up (Table e3, Supplementary Appendix).

The magnitude of the beneficial effect of therapeutic anticoagulation on the primary outcome was maintained in all subgroup analysis (Table e9, Supplementary Appendix).

### Safety

Serious adverse events were reported in 12 patients (16%) in therapeutic anticoagulation group, 15 patients (18.8%) in prophylactic anticoagulation group, 18 patients (22.8%) in therapeutic anticoagulation plus tocilizumab group, and 21 patients (28.4%) in prophylactic anticoagulation plus tocilizumab group (P=0.277) (Table 3). Septic shock was diagnosed in 8 (10.7%) patients in therapeutic anticoagulation group, in 13 (16.3%) patients in prophylactic anticoagulation group, in 18 patients (22.8%) in therapeutic anticoagulation plus tocilizumab group, and in 21 patients (28.4%) in prophylactic anticoagulation plus tocilizumab group (P=0.036).

**Table 3.** Safety.

| Events | Therapeutic AC<br>n=75 | Prophylactic AC<br>n=80 | Therapeutic AC +<br>Tocilizumab<br>n=79 | Prophylactic AC+<br>Tocilizumab<br>n=74 | p |
| --- | --- | --- | --- | --- | --- |
| Total (%) | 12 (16.0%) | 15 (18.8%) | 18 (22.8%) | 21 (28.4%) | .277<sup>a</sup> |
| Major bleeding* | 5 (6.7%) | 1 (1.3%) | 4 (5.0%) | 0 (0.0%) | .029<sup>b</sup> |
| HIT | 0 (0.0%) | 1 (1.3%) | 0 (0.0%) | 0 (0.0%) | .44<sup>b</sup> |
| Septic shock | 8 (10.7%) | 13 (16.3%) | 18 (22.8%) | 21 (28.4%) | .036<sup>c</sup> |

\*Major bleeding: clinically overt bleeding accompanied by a decrease in the Hb level of $\geq 2$ g/dL or transfusion of $\geq 2$ units of packed red cells, occurring at a critical site, or resulting in death. HIT: Heparin induced thrombocytopenia; AC: anticoagulation. a: Pearson's chi-squared test; b: likelihood-ratio test; c: Kruskal-Wallis test.

Other adverse events of especial interest are provided in the Table e10 (Supplementary Appendix).

The distribution of patients’ scores on the seven-level ordinal scale at 7 and 30 days is showed in the Figure e1 (Supplementary Appendix).

## Discussion

In this multicenter randomized trial, therapeutic anticoagulation was the best strategy in terms of improved clinical status at 30-days and reduced mortality in hospitalized patients with severe COVID-19. This is the first trial analyzing the combination of different regimens of heparin with or without tocilizumab in a particular population of severe ill patients admitted in tertiary health care facilities. Understanding the various phenotypes of COVID-19 and how they may require different treatments and may result in distinct therapeutic responses based on individual data is essential to better define early strategies to reduce complications and mortality. The benefit of therapeutic heparin might be due to, despite anticoagulant effect, anti-inflammatory and immunomodulatory effects in preventing thrombosis and reducing inflammation in the early stages of COVID-19 infection.[27]

We included patients with severe COVID-19 within the first 10 days of clinical course. At randomization, most patients, despite of presenting the severe form of disease, were still at the emergency department, because of limited intensive care unit bed availability in Brazil during the pandemics peak.

Several high-quality randomized trials have recently been published to define the role of therapeutic dose anticoagulation for hospitalized COVID-19 patients.^6-11^ Relevant studies showed that therapeutic heparin adds benefits to patients with moderate disease and not requiring ICU level care. The ATTACC, ACTIV-4a, and REMAP-CAP platform trials demonstrated reduction in organ support-free days with therapeutic dose heparin in non-critically ill patients, with a treatment benefit more apparent in patients with high D-dimer.[28] In contrast, the largest recently published trial, did not observe any statically significant differences concerning the composite primary outcome when comparing prophylactic-dose heparin, therapeutic-dose enoxaparin, or therapeutic-dose apixaban.[29] The authors of this multiplatform group have also published their findings regarding treatment of critically ill patients with therapeutic dose heparin. The trial was stopped prematurely due to futility despite having enrolled more than 1000 patients.[30]

ACTION trial showed that therapeutic dose of rivaroxaban or enoxaparin followed by rivaroxaban to day 30 did not significantly reduced clinical outcomes (time to death, duration of hospitalization or duration of supplemental oxygen to day 30), while it increased bleeding compared with prophylactic anticoagulation.[10] The INSPIRATION trial evaluated critically ill patients with COVID-19 and found that intermediate-dose anticoagulant regimen did not reduced the incidence of organ support-free days or mortality compared to standard prophylactic-dose of subcutaneous UFH or subcutaneous enoxaparin.[11] The HEP-COVID trial demonstrated that among hospitalized COVID-19 patients with a D-dimer level ≥4 times the upper limit of normal, therapeutic dose heparin improved outcomes without increasing risk for major bleeding. The observed effect was evident among the non-ICU patients, whereas no significant effect was observed among ICU patients.[12] The RAPID trial did not show benefit of therapeutic heparin versus prophylactic heparin to reduce the composite primary outcome of death, need for invasive mechanical ventilation and admission to an ICU in moderately-ill patients with elevated D-dimer levels.[13]

Various underlying factors could account for the observed advantages of heparin in our study. Most heparin trials were multicenter and multinational, enrolling patients with diverse disease phenotypes and distinct responses to different therapies. Individual characteristics of patients, such as racial background, viremia, inflammatory, and thrombotic mechanisms might determine heparin benefits in COVID-19. Our study included exclusively severely ill patients who required intensive critical care, most using non-invasive mechanical ventilation or invasive mechanical ventilation following hospitalization within a recent timeframe. Heparin was administered in most patients within a period of two days following their admission within 7 days of the onset of symptoms. Most patients were randomized during their admission in the emergency ward, in the initial stage of COVID-19. Probably, therapeutic heparin avoided thromboinflammation before the development of an irreversible hyperinflammatory state and cytokine storm, which may occur in late-stage of disease.[31] Non-anticoagulant mechanisms underlying treatment of COVID-19 patients with heparin include: (I) inhibition of heparanase activity, preventing endothelial leakage; (II) neutralization of chemokines and cytokines; (III) interference with leukocyte trafficking; (IV) reduction of viral cellular entry, and (V) neutralization of extracellular cytotoxic histones.[32]

In previous studies, moderate disease severity was defined as hospitalization for COVID-19 without the need for ICU level care. In ACTIV-4a, in which investigators found that ICU-level care was challenging to define during the pandemic, needing of organ support, was used to define ICU level care. Patients who were admitted to an ICU but without receiving qualifying organ support were considered moderately ill.^10,11^ The lack of consensus on definitions of disease severity during pandemics has led to conflicting results in studies evaluating the efficacy of various therapeutic interventions, inhibiting guidelines to establish clinical decision-making and accurate comparisons between studies.

As part of our investigation, tocilizumab with prophylactic or therapeutic anticoagulation did not result in clinical improvement in severe manifestations of disease and led to a higher incidence of septic shock and increased mortality rates.

A recent meta-analysis included 15 randomized studies involving a total of 923 patients with COVID-19 who received one or two doses of IL-6 inhibitors, with 13 trials using tocilizumab and 2 trials using sarilumab.[20] IL-6 inhibitors administration in patients with moderate or severe COVID-19 pneumonia reduced longest follow-up mortality. Furthermore, it was associated with reduction in 28/30-day mortality, need for intubation and clinical worsening, without increasing adverse effects such as septic shock. It should be noted, however, that the positive effects of tocilizumab on mortality were primarily influenced by the results of the RECOVERY trial.[25] Within the context of our investigation, tocilizumab resulted in higher incidence of septic shock as compared to the groups that did not receive tocilizumab. The absence of benefits regarding to the primary outcome in patients assigned to tocilizumab with therapeutic heparin may be attributed to the fact that our study involved critically ill patients admitted during the pandemic, with a 15% incidence of septic shock, which is considerably higher than the 1-5% of incidence reported in previous trials.[17] Recent publications have also revealed significant information regarding the heightened risk of associates infection in COVID-19 patients who have been treated with tocilizumab.[33-35] Although it is still a subject of debate and different perspectives[36], there is no consensus on the issue regarding the safety of tocilizumab.

Patients included in this study presented elevated levels of severity biomarkers including CRP, D-dimer, and IL-6 at baseline evaluation, indicating a heightened risk of complications and mortality, and could potentially serve as predictors of the effectiveness of anti-inflammatory therapies.[37] In this case, therapeutic heparin reduced significantly CRP during 7 days of follow-up, suggesting an anti-inflammatory effect as an important therapeutic mechanism in COVID-19.

## Limitations

This trial has several limitations. A double-blind study in Brazil was not feasible due to logistical constraints during the pandemics, resulting in lack of blinding. However, investigators were not taking care directly of the patients, and outcome assessors were not aware of the treatment arms. The primary outcome has important flaws, including sensitivity to differences in local clinical practice, which might limit the generalizability of our findings, but at the same time it comprises important endpoints such as escalating of inpatient care and mortality. Lastly, our investigation focused on a specific COVID-19 population in the early stages of the disease requiring at least 4 L/min of oxygen, which restricts the applicability of our findings to other groups of patients.

## Conclusions

In severe COVID-19 patients, therapeutic anticoagulation with heparin increased the proportion of recovery in 30 days. Even if therapeutic anticoagulation increased bleeding, it was associated with a reduced overall mortality. Tocilizumab did not provide additional benefits to heparin in COVID-19 patients.

## Data Availability

Data are available under requestion after publication.

## List of abbreviations

COVID-19: Coronavirus disease 2019
IL-6: interleukin 6
SARS-CoV-2: Severe Acute Respiratory Syndrome due to Coronavirus-2
RT-PCR: real-time polymerase chain reaction
UFH: unfractionated heparin
aPTT: activated partial thromboplastin time
CRP: C-reactive protein
NT-proBNP: N-terminal pro B-type natriuretic peptide
WHO: World Health Organization
AKIN: Acute Kidney Injury Network
ICU: Intensive Care Unit
ITT: intention-to-treat
CI: confidence interval
SD: standard deviation

## Declarations

### Ethics approval and consent to participate

The protocol and informed consent were approved by the ethics committees of Hospital das Clínicas, Faculdade de Medicina, Universidade de São Paulo (approval number 4.119.201, 28 June, 2020) and each participating site.

### Consent for publication

Not applicable.

## Availability of data and materials

*Data available:* Yes.

*Data types:* Deidentified participant data.

*How to access data:* Patient-level data and full dataset and statistical code will be available upon reasonable request to the corresponding author.

*When available:* With publication.

## Competing interests

The authors declare no competing interests.

## Funding

The trial was funded by the *Conselho Nacional de Desenvolvimento Científico e Tecnológico (CNPq Grant Number 317640/2021-6)*, an institution linked to the Ministry of Science and Technology, Brazil.

## Authors’ contributions

Drs Ribeiro and Hajjar had full access to all of the data in the study and take responsibility for the integrity of the data and the accuracy of the data analysis.

*Concept and design:* Ribeiro, Hajjar, Vieira, Huemer, Landoni, Kalil Filho. *Acquisition, analysis, or interpretation of data:* Lacerda, Rizk, Costa, Andrade, Melo, Santos Neto, González, Reis, Costa, Machado, Ribeiro, Vieira, Huemer, Silva, Park, Fukushima, Guimarães Junior, Cota, Kawahara, Barros, Muller, Nakada, Pinto, Bergamin, Santos, Figueira, Paula, Kalil Filho.

*Critical revision of the manuscript for important intellectual content:* Ribeiro, Hajjar, Quintão, Vieira, Huemer, Landoni, Kalil Filho.

*Statistical analysis:* Fukushima.

*Administrative, technical, or material support:* Vieira, Huemer.

*Supervision:* Hajjar, Landoni, Kalil Filho.

## Acknowledgements

None

## Notes

### Competing Interest Statement

The authors have declared no competing interest.

### Clinical Trial

Clinicaltrials.gov NCT04600141. Registered October 22, 2020.

### Author Declarations

The protocol and informed written consent were approved by the ethics committees of Hospital das Clínicas, Faculdade de Medicina, Universidade de São Paulo (approval number 4.119.201, 28 June, 2020) and each participating site.

